# Clinical evaluation of the SD Biosensor saliva antigen rapid test with symptomatic and asymptomatic, non-hospitalized patients

**DOI:** 10.1101/2021.04.21.21255865

**Authors:** Zsofia Igloi, Jans Velzing, Robin Huisman, Corine Geurtsvankessel, Anoushka Comvalius, Janko van Beek, Roel Ensing, Timo Boelsums, Marion Koopmans, Richard Molenkamp

## Abstract

Performance of saliva antigen rapid test was evaluated in non-hospitalized patients, with or without symptoms. Overall sensitivity was 66.1% compared to RT-PCR in saliva. Using cycle threshold <30 cutoff or virus culture as reference, sensitivity increased to 88.6% or 96.7% respectively. Specificity was 99.6%.

## Introduction

In spite of having multiple vaccines available (1), the COVID-19 pandemic is far from over and diagnostic capacities remain an essential pillar of the pandemic response. To broaden access to testing, self / home testing approaches are explored worldwide although there is considerable debate on the reliability of self-testing. In order to make self / home testing available, clear testing guides need to be developed suitable for all ages and education levels. Furthermore besides a few (2-4), most antigen rapid tests (Ag RDTs) have currently only been validated using nasopharyngeal swabs which arguably can only be executed by trained health-care professionals and are less suited for self-testing. Here we report results of a clinical evaluation of a not yet marketed antigen rapid test using saliva.

## Methods

### Testing site, testing procedures, population and patient recruitment

The study was carried out at an XL testing location in Rotterdam - Rijnmond (Rotterdam is the second largest city in the Netherlands) which is by appointment only. Persons with any respiratory symptoms, or persons that have been contacts of confirmed cases regardless of symptoms are eligible for a free of charge SARS-CoV-2 test. Vulnerable persons (ie elderly or chronic condition) and priority groups (ie teachers, healthcare workers) are tested by reverse transcriptase polymerase chain reaction (RT-PCR), everyone else is tested by the SARS-CoV-2 Rapid Antigen Test (Distributed by Roche (SD Biosensor)) using a nasopharyngeal swab. In this study persons eligible for a rapid antigen test were enrolled. The SARS-CoV-2 Rapid Antigen Test-Standard Q COVID-19 Ag Saliva-Research use only (Lot number QCO9021001; expiry date 04-01-2023) was provided by SD Biosensor (http://www.sdbiosensor.com/xe/) but at time of writing this rapid antigen test was not yet available on the market.

At the entrance of the testing site all people over 18 years of age were approached for inclusion and enrolled following verbal informed consent. Study participants were also asked to fill in a questionnaire stating age, sex, date of symptom onset. Recommended time of ≥30mins since last meal, drink or cigarette if applicable was also recorded. The study was carried out for 4 weeks to reach an ideally total of 1000 inclusions (inclusion was maximized to 50/day due to logistics). Study was terminated just prior to reaching the target at 816 inclusions for logistic reasons.

### Specimen collection, testing and culture procedures

Saliva was collected based on instructions of use by the test provider (nasal discharge and cough discharge drooled into collection device) using the Zeesan Saliva RNA collection kit without preservation medium. (http://www.zeesandx.com/coronavirus/1075.html). All saliva samples were tested for SARS-CoV-2 RNA using the cobas ® SARS-CoV-2 RT-PCR test on the COBAS6800 (Roche diagnostics). Genome copies/ml were calculated based on an in house established standard curve (5). All RT-PCR positive saliva samples were inoculated onto Vero cells following dilution with universal transport media (VTM) (HiViral; HiMedia Laboratories PVT, Ltd., https://www.himedialabs.com) to 6ml volume, filtration with 45µm bacterial filter and mixing with FBS: 1080ul saliva+VTM with 720ul FBS prior to freezing at -80C for a maximum of two weeks. Samples were cultured for a maximum of 14 days or until cytopathic effect (CPE) was visible. The presence of SARS-CoV-2 was confirmed by immunofluorescence, using a rabbit polyclonal antibody targeting SARS CoV-2 nucleocapsid protein (Sino Biological Inc.).

### Data analysis

Data from the Ag RDTs, RT-PCR, virus culture, and clinical questionnaire were merged using Microsoft Access (http://www.microsoft.com), and data analysis was performed using Microsoft excel and R software version 4.0.2 (https://www.r-project.org). Sensitivity and specificity of Ag RDT were calculated in relation to the RT-PCR, nasopharyngeal Ag RDT and virus culture results. Clopper-Pearson analysis was used to determine confidence intervals of proportions. Two sample t-test was used to define significance of difference between means.

### Ethical clearance

The medical research ethics committee (MREC) of Erasmus Medical center decided the study was not subject to the Medical Research Involving Human Subjects Act (WMO) and did not require full review by an accredited MREC (protocol number MEC-2021-0083).

## Results

Most people presenting for testing had symptoms (70.5%, 556/789) with recent onset (median 2 days post onset) (Table 1). Participants (n=789) had a median age of 37 years and almost equal proportion of males and female (male, 50.6%). Sensitivity of the saliva Ag RDT was analyzed in three ways : i, compared to saliva RT-PCR; ii, compared to nasopharyngeal Ag RDT and iii, compared to virus culture (Table 2). Overall sensitivity ranged between 66.1%-96.7% depending on the reference method used. When PCR ct <30 (E gene copy/ml 2.17E+05) cutoff was used sensitivity increased to 88.6% (CI95% 75.4 - 96.2). Overall performance of saliva Ag RDT was inferior to nasopharyngeal Ag RDT compared to saliva RT-PCR without or with ct <30 cutoff (66.1% vs 79.0% and 88.6% vs 90.9%). Analysis by days post onset did not result in higher sensitivities. When breaking down the data for symptomatic and asymptomatic individuals, sensitivity ranged from 82.7% to 60% respectively, however the total number of RT-PCR positive asymptomatic participants was very low (n=10). Specificity was calculated compared to saliva RT-PCR and nasopharyngeal Ag RDT and was comparable to previous findings using nasopharyngeal Ag RDT (between 99.3% and 99.6%) (5).

**Table 1.**
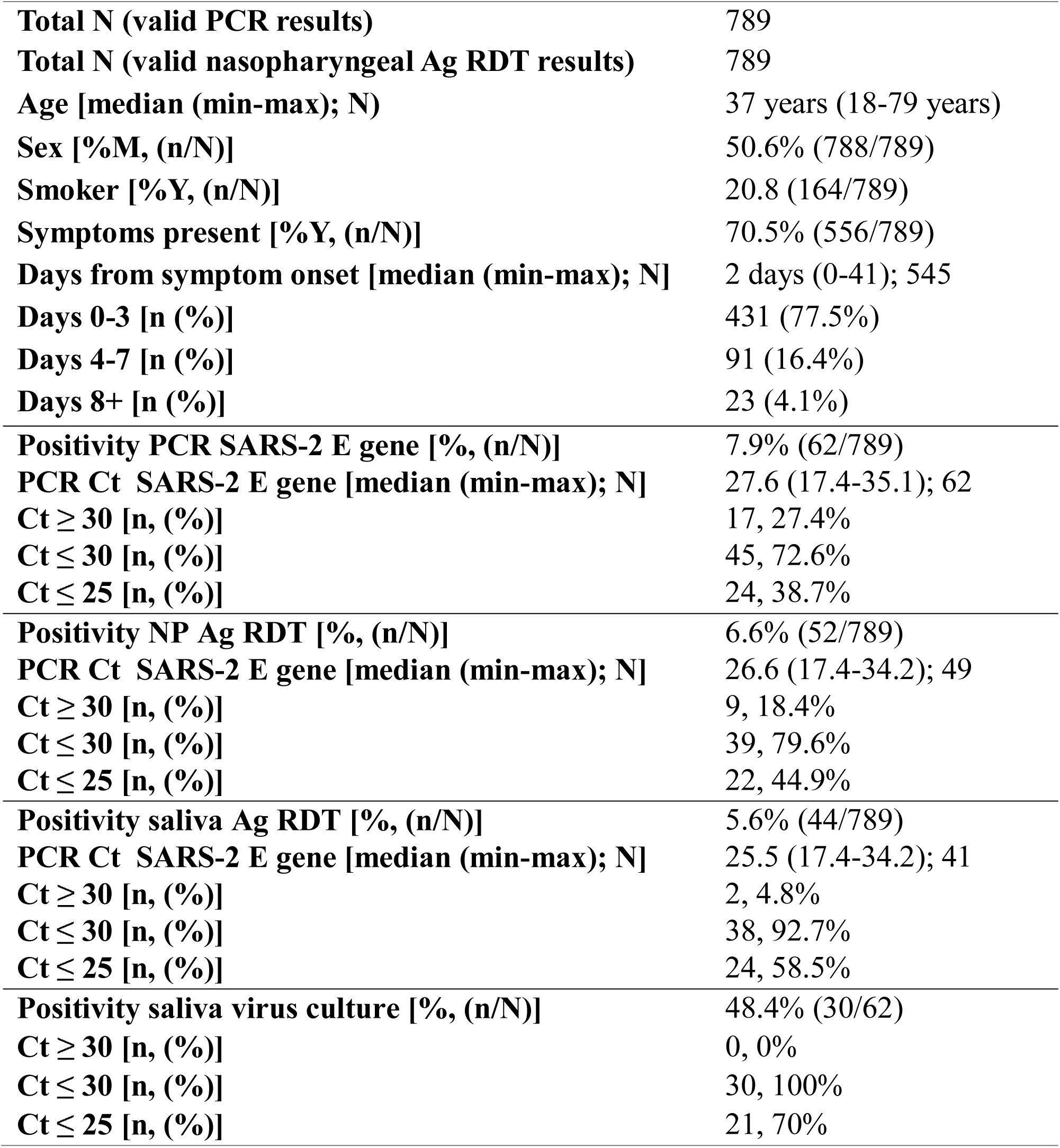
Characteristics of the study population. Data of all included people in the study were analyzed by basic demographics, smoking status, date of disease onset, PCR and nasopharyngeal Ag RDT results also by ct values.

|  |  |
| --- | --- |
| **Total N (valid PCR results)** | 789 |
| **Total N (valid nasopharyngeal Ag RDT results)** | 789 |
| **Age [median (min-max); N]** | 37 years (18-79 years) |
| **Sex [%M, (n/N)]** | 50.6% (788/789) |
| **Smoker [%Y, (n/N)]** | 20.8 (164/789) |
| **Symptoms present [%Y, (n/N)]** | 70.5% (556/789) |
| **Days from symptom onset [median (min-max); N]** | 2 days (0-41); 545 |
| **Days 0-3 [n (%)]** | 431 (77.5%) |
| **Days 4-7 [n (%)]** | 91 (16.4%) |
| **Days 8+ [n (%)]** | 23 (4.1%) |
| **Positivity PCR SARS-2 E gene [%, (n/N)]** | 7.9% (62/789) |
| **PCR Ct SARS-2 E gene [median (min-max); N]** | 27.6 (17.4-35.1); 62 |
| **Ct ≥ 30 [n, (%)]** | 17, 27.4% |
| **Ct ≤ 30 [n, (%)]** | 45, 72.6% |
| **Ct ≤ 25 [n, (%)]** | 24, 38.7% |
| **Positivity NP Ag RDT [%, (n/N)]** | 6.6% (52/789) |
| **PCR Ct SARS-2 E gene [median (min-max); N]** | 26.6 (17.4-34.2); 49 |
| **Ct ≥ 30 [n, (%)]** | 9, 18.4% |
| **Ct ≤ 30 [n, (%)]** | 39, 79.6% |
| **Ct ≤ 25 [n, (%)]** | 22, 44.9% |
| **Positivity saliva Ag RDT [%, (n/N)]** | 5.6% (44/789) |
| **PCR Ct SARS-2 E gene [median (min-max); N]** | 25.5 (17.4-34.2); 41 |
| **Ct ≥ 30 [n, (%)]** | 2, 4.8% |
| **Ct ≤ 30 [n, (%)]** | 38, 92.7% |
| **Ct ≤ 25 [n, (%)]** | 24, 58.5% |
| **Positivity saliva virus culture [%, (n/N)]** | 48.4% (30/62) |
| **Ct ≥ 30 [n, (%)]** | 0, 0% |
| **Ct ≤ 30 [n, (%)]** | 30, 100% |
| **Ct ≤ 25 [n, (%)]** | 21, 70% |

**Table 2.** Sensitivity and specificities of both nasopharyngeal and saliva antigen RDT compared to RT-PCR and saliva antigen RDT compared to nasopharyngeal RDT. Overall and stratified sensitivity and specificity of the saliva antigen RDT compared to RT PCR and the nasopharyngeal antigen RDT. Results were also analyzed by days post onset and symptom status. Positive and negative predictive values were calculated using 7.9% prevalence setting.

| Compared to saliva RT-PCR |  |  |  |  |  |
| --- | --- | --- | --- | --- | --- |
| NP RDT | 0-3 days post onset | 0-7 days post onset | Overall | Overall-symptomatic | Overall-asymptomatic |
| **Clinical Sensitivity, % (CI95%), N** | 79.2%<br>(65.0- 89.5) (666) | 78.7%<br>(66.3-88.1) (764) | 79.0%<br>(66.8 88.3)(789) | 82.7%<br>(69.7- 91.8) (556) | 60.0%<br>(26.2-87.9) (233) |
| **<ct 30, Clinical Sensitivity%, (CI95%), N** | 90.0%<br>(76.3-97.2) (428) | 90.5%<br>(77.4-97.3) (747) | 90.9 %<br>(78.3-97.5) (770) | 91.2%<br>(76.3-98.1) (432) | 83.3%<br>(35.9-99.6) (228) |
| **Clinical Specificity, % (CI95%)** | 99.5%<br>(98.6- 99.9) | 99.58%<br>(98.8-99.9) | 99.6%<br>(98.8-99.9) | 99.6%<br>(98.6- 99.9) | 99.6%<br>(97.5-99.9) |
| **NPV, % (CI95%)** | 98.2%<br>(97.0-99.0) | 98.2%<br>(97.1-98.9) | 98.2%<br>(97.2- 98.9) | 98.5%<br>(97.4-99.2) | 96.7%<br>(93.2-98.4) |
| **PPV, % (CI95%)** | 93.4%<br>(81.8-97.8) | 94.1%<br>(83.6-98.0) | 94.3%<br>(84.1-98.1) | 94.7%<br>(81.7-98.6) | 92.0%<br>(60.4-98.9) |
| **Saliva RDT** |  |  |  |  |  |
| **Clinical Sensitivity, % (CI95%), N** | 68.5%<br>(54.5-80.5) (673) | 65.0%<br>(51.6-76.9) (763) | 66.1%<br>(52.9-77.6) (789) | 69.2%<br>(54.9-81.3) (556) | 50.0%<br>(18.7-81.3) (233) |
| **<ct 30, Clinical Sensitivity, % (CI95%), N** | 87.5%<br>(73.2-95.8) (428) | 88.1%<br>(74.4-96.0) (747) | 88.6%<br>(75.4-96.2) (770) | 88.2%<br>(72.6-96.7) (432) | 83.3%<br>(35.9-99.6) (228) |
| **Clinical Specificity, % (CI95%)** | 99.7%<br>(98.8-99.9) | 99.6%<br>(98.8-99.9) | 99.6%<br>(98.8-99.9) | 99.8%<br>(98.9- 99.9) | 99.1%<br>(96.8-9.9) |
| **NPV, % (CI95%)** | 97.4%<br>(96.1-98.2) | 97.1%<br>(95.9%-97.9) | 97.2%<br>(96.0-97.9) | 97.4%<br>(96.2-98.3) | 95.9%<br>(92.6%-97.7) |
| **PPV, % (CI95%)** | 94.8%<br>(81.9-98.7) | 92.9%<br>(80.7-97.6) | 93.2%<br>(81.4- 97.7) | 96.8%<br>(80.7-99.5) | 82.7%<br>(51.3-95.6) |

| Compared to nasopharyngeal Ag RDT |  |  |  |  |  |
| --- | --- | --- | --- | --- | --- |
| Saliva RDT | 0-3 days post onset | 0-7 days post onset | Overall | Overall-symptomatic | Overall-asymptomatic |
| Clinical Sensitivity, % (CI95%), N | 74.5% (59.7-86.1) (671) | 74.0% (59.7-85.4) (761) | 75.0% (61.1-85.9) (789) | 75.6% (60.5-87.1) (556) | 50.0% (18.7-81.3) (233) |
| <ct 30, Clinical Sensitivity,% (CI95%), N | 84.6 (69.5-94.2) (425) | 85.4% (70.8-94.4) (748) | 86.1% (72.1-94.7) (770) | 84.9% (68.1-94.9) (432) | 83.3% (35.9-99.6) (228) |
| Clinical Specificity, % (CI95%), | 99.4% (98.4-99.8) | 99.3% (98.4-99.8) | 99.3% (98.4-99.8) | 99.4% (98.3-99.9) | 99.1% (96.8-99.9) |
| NPV, % (CI95%) | 97.8% (96.5- 98.7) | 97.8% (96.5-98.6) | 97.9% (96.7-98.7) | 97.9% (96.6-98.8) | 97.6% (92.6-99.2) |
| PPV, % (CI95%) | 90.9% (78.8-96.4) | 90.1% (78.9-95.7) | 90.5% (79.6-95.8) | 91.7% (77.9-97.2) | 87.4% (61.7-96.8) |

All PCR positive saliva samples were used for cell culture and approximately half (48.4%, 30/62) resulted in a positive cell culture (mean ct 24.2 of positive samples) (Figure 1.). Almost all samples with a positive culture results were detected either by nasopharyngeal RDT (90%, 27/30) or by saliva RDT (97%, 29/30). Only one sample was not detected by both RDTs and demonstrated a relatively high ct-value (ct 29). The corresponding patient was most likely in the very early phase of onset (day 0 of symptoms). Culture negative samples (51.6%, 32/62) had higher ct values (mean ct 30.2 (p= 0.001). Results of the two RDTs were partially in concordance as 10/32 were not detected by nasopharyngeal RDT or by saliva RDT. The culture negative samples which were not detected by the saliva Ag RDT only were in the high ct value range (median ct 31.5).

**Figure 1.**
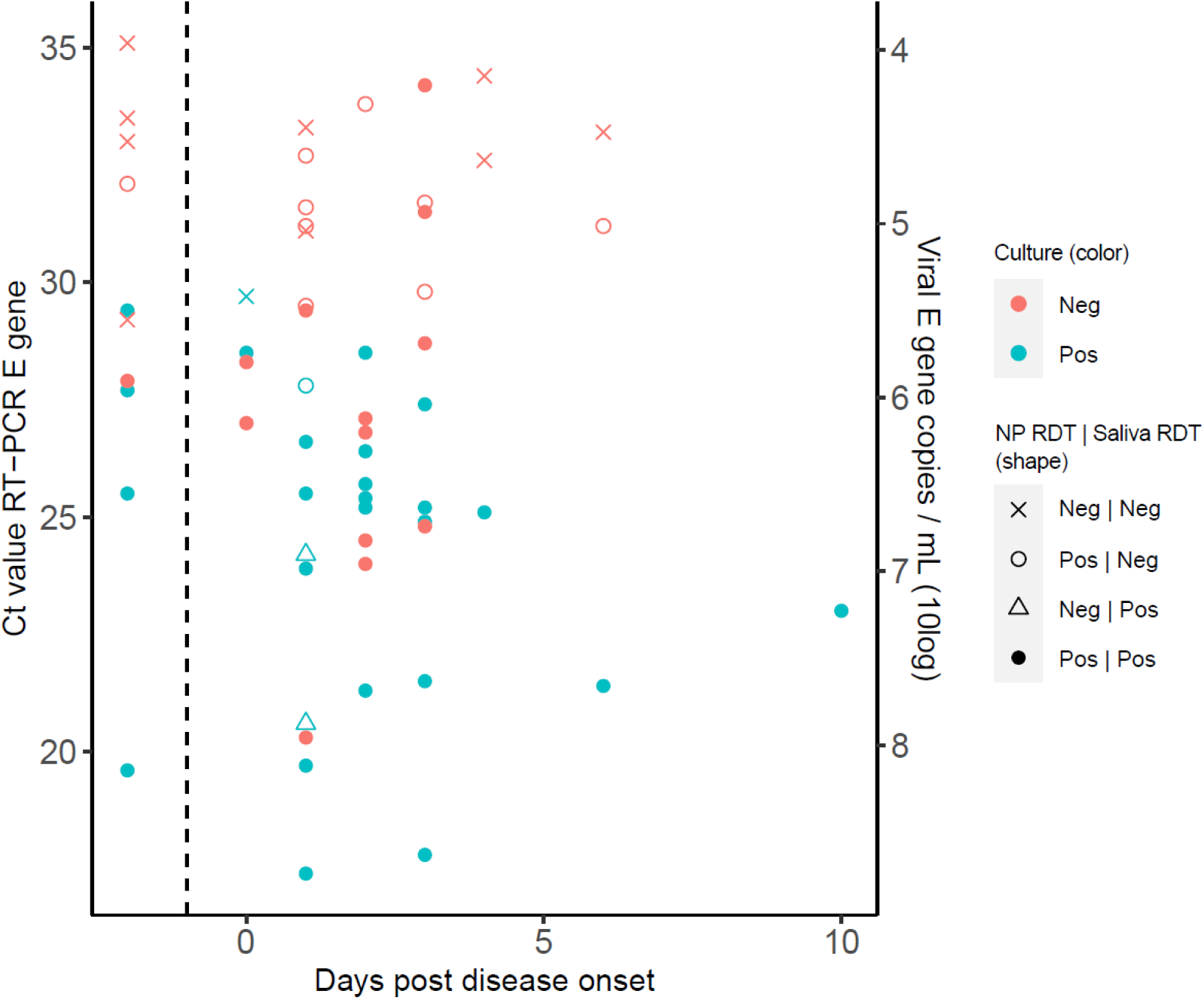
Cycle thresholds and genome copies of positive samples in relation to days since symptom onset, saliva and nasopharyngeal Ag RDT positivity, and culture outcomes of participation with both symptomatic and asymptomatic patient (n = 59).). Data points shown on the left side of the dashed bar are from asymptomatic individuals. NP, nasopharyngeal swab; Ag RDT, antigen rapid detection test; Ct, cycle threshold; E gene, envelope gene; Neg, negative; Pos, positive; RT-PCR, reverse transcription PCR.

## Discussion

There is very limited data available on the performance of saliva based Ag RDTs (6, 7) and those that are available show below optimal results. The published studies report sensitivities of 63% and 24% compared to nasopharyngeal or nasal swab utilizing Ag RDTs. In our study we have found that people with high viral load (ct<30 cutoff) and early phase of disease could be detected with a saliva-based antigen RDT with relatively high sensitivity (88.6% CI95% 75.4 - 96.2). Furthermore, the majority of presumed infectious individuals (based on cell-culture positivity in saliva) could be detected (96.7%). However there is still much to learn about viral kinetics, the role of SARS-CoV-2 specific antibodies in saliva and viral load correlation to disease severity.

In this study, nasopharyngeal Ag RDT was used as default diagnostic method and also as reference method for the saliva Ag RDT and demonstrated very comparable sensitivity/specificity to our previous evaluation (5). Using saliva RDT, despite lower general sensitivity compared to both saliva RT PCR and nasopharyngeal RDT, most infectious individuals could be detected, however this result was not compared to nasopharyngeal virus culture. Saliva is a complex material shown to have comparable or slightly lower viral load to nasopharyngeal sampling sites (8), it also contains SARS-2 specific antibodies (9) and different viral kinetics compared to nasopharynx which might explain the lower performance in the tested population. We have evaluated this test on non-hospitalized patients; a study found correlation of more severe disease with higher viral load in saliva (10).

Although our study is limited in the suboptimal number of positive samples it provides promise for further exploration of saliva based Ag RDTs as they could be used for self/home testing. We did not investigate acceptance of saliva vs other invasive sample types but one could argue that collection of saliva is easier to perform and causes minimal discomfort. In addition due to the ease of use, variation in sample collection between individuals might be low, limiting the risk of improper sample collection. Limited studies on usability showed relatively high sensitivity and low false negativity related to self-testing (11), however comprehensive studies are still mostly lacking.

Overall, the potential benefits of saliva Ag RDT, could outweigh the drawbacks in a comprehensive testing strategy, especially for home/self testing and in vulnerable populations like elderly, disabled or children where in intrusive testing is either not possible or causes unnecessary stress.

## Data Availability

all data included in the manuscript

## Acknowledgement

Testing was carried out at the Ahoy XL testing centre in Rotterdam, the Netherlands, where numerous people contributed to the success of this project. We would like to thank all study participants, employees of the GGD Rotterdam - Rijmond especially all laboratory coordinators, namely Jelena Pajic, Lars Kortenhorst, Niekeline Kimmel, Ono Bestman, Bert de Valk, Sylviana Anthony for coordination and support and the laboratory workers who carried out rapid antigen tests.

## Funding

The SARS-CoV-2 Rapid Antigen Test-Standard Q COVID-19 Ag Saliva-Research use only (Lot number QCO9021001; expiry date 04-01-2023) was provided by SD Biosensor No other funding was received.

## Author Bio

Zsofia Igloi is a virologist/public health microbiologist working at the Erasmus Medical Centre Viroscience department and has a primary interest in arbo- and emerging viruses, diagnostics and public health implications.

## Notes

### Competing Interest Statement

The authors have declared no competing interest.

### Funding Statement

no external funding received

